# Evaluation of a Multidimensional Assessment Tool to Simultaneously Determine Physical Activity, Nutritional and Quality of Life Status in Cancer Patients

**DOI:** 10.1101/2025.10.05.25337352

**Authors:** Sebastian Theurich, Eva Kerschbaum, Annika Tomanek, Christine Welker, Timo Niels, Nicole Erickson, Hansjörg Baurecht, Nora Zoth, Michael Leitzmann, Freerk T. Baumann

**Affiliations:** Department of Medicine III, LMU University Hospital Munich, Munich, Germany; Cancer- and Immunometabolism Research Group, Gene Center, LMU Munich, Germany; Comprehensive Cancer Center Munich (CCCM), LMU Munich, Munich, Germany; German Cancer Consortium (DKTK), Munich Site, and German Cancer Research Center, Heidelberg, Germany; Department I of Internal Medicine, Center of Integrated Oncology Aachen Bonn Cologne Düsseldorf, University Hospital of Cologne, Cologne, Germany; Faculty of Medicine, Institute of Epidemiology and Preventive Medicine, University of Regensburg, Regensburg, Germany

**Author notes:** **Corresponding Authors:** Sebastian Theurich, LMU University Hospital Munich, Department of Medicine III And Freerk T. Baumann, University Hospital Cologne, Center for Integrated Oncology (CIO). contributed equally (Sebastian Theurich, Eva Kerschbaum).

**Keywords:** Cancer, Nutrition Counselling, Physical Exercise Intervention, Outcomes Research

## Abstract

**Background:** In cancer patients, the assessment of malnutrition, muscular and neurological deficits and quality of life are usually performed independently from each other, which neglects mutual interactions, and thus, lowers the potential of supportive interventions. Therefore, we developed the “resource-oriented needs assessment (RoBa)” as a multidimensional assessment battery that simultaneously captures i) the nutritional status, ii) physical fitness and iii) the psycho-oncological status based on four validated tools or questionnaires in each domain. In a prospective, multicenter pilot study, we evaluated the feasibility and reliability of the RoBa score in real-world cancer care.

**Methods:** Consecutive cancer patients from clinical routine care at two university cancer centers were prospectively included and underwent all twelve assessments. The physical fitness domain contained the handgrip strength test, spiro-ergometry, one-leg stand test, and seven-day accelerometry. Assessments of the nutritional status domain covered the Patient-Generated Subjective Global Assessment (PG-SGA), Body-Mass-Index (BMI), Bioelectrical Impedance Analysis (BIA), and the modified Glasgow-Prognostic-Score (mGPS). The quality-of-life status was assessed by the Multidimensional Fatigue-Inventary (MFI-20), Functional Assessment of Cancer Therapy/Gynecologic Oncology Group – Neurotoxicity (FACT/GOG-Ntx), Hospital Anxiety and Depression Scale (HADS), and the EORTC QLQ-C30 questionnaire. The results of each individual assessment were scored 0 (no needs), 1 (moderate needs), or 2 (severe needs) based on published cut-off values of each assessment or international guidelines. Individual scores were summarized to domain and total RoBa scores. Correlation analyses were performed with individual, domain and total RoBa score data.

**Results:** Between 2022 and 2024 a total of 62 consecutive cancer patients (GI cancer (40.3%), non-GI cancer (59.7%) were included in to the study at two academic cancer centers in Germany. From all three RoBa domains, quality of life individual scores showed the lowest deviation from the respective domain score. Within the three domains, the two strongest correlations of individual scores with the domain score were seen for PG-SGA (r = 0.64) and BIA (r = 0.54) within the nutrition domain, spiroergometry (r = 0.85) and accelerometry (r = 0.55) within the physical fitness domain, and MFI-20 (r = 0.81) and EORTC-Q30 (r = 0.74). With regard to the total RoBa score, high correlations of each domain score were also observed: nutrition domain (r = 0.59), physical fitness (r = 0.69), and quality of life (r = 0.65). Subgroup analyses of GI versus non-GI-cancer patients revealed differences in the nutritional domain scorings

**Conclusion:** The RoBa scoring system proved feasible and strong correlations to the published assessment evaluations but also within the domain and total RoBa scoring system were observed in this pilot study. The score yielded consistent classifications regardless of the tested tumor entity, supporting its implementation for integrated estimation of support needs in nutrition, exercise, and psycho-oncology. Further research in larger cohorts is warranted.

## Introduction

Cancer patients are at risk to develop malnutrition, sarcopenia, or a low clinical performance status during their treatment, or already show these symptoms at diagnosis. Not only are these well-established risk factors that impact on cancer treatment efficacy but also mediate or facilitate therapy-associated toxicities. In addition, the nutritional status, physical fitness and mobility, as well as the mental and psychological status are closely connected, and mutually interfere. Surprisingly, there is currently no clinical standard to screen and assess all these aspects in a multidimensional manner. Instead, clinically established and standardized tools in clinical nutrition and physical activity assess only one dimension at the same time. Here, the Malnutrition Universal Screening Tool (MUST), Nutritional Risk Screening (NRS-2002), and Patient-Generated Subjective Global Assessment (PG-SGA) are widely used to detect malnutrition risk [28]. If indicated, guidelines recommend extended diagnostics including dietary intake, body composition, performance capacity, and systemic inflammation, as these parameters correlate with patient outcomes [3, 29]. Methods such as bioelectrical impedance analysis (BIA), dual-energy X-ray absorptiometry (DEXA), and computed tomography (CT) allow for the assessment of muscle mass, while C-reactive protein (CRP), albumin, and the modified Glasgow Prognostic Score (mGPS) provide markers of inflammation and catabolism [2, 29].

Physical activity is another critical factor. ASCO guidelines recommend endurance and resistance training during cancer therapy when medically feasible [30]. Activity levels can be quantified using metabolic equivalents (METs), moderate-to-vigorous physical activity (mvpa), questionnaires, or accelerometers [31–34]. Complementary indicators include handgrip strength, balance tests, and spiroergometry for cardiorespiratory fitness [35].

Quality of life is commonly assessed with standardized questionnaires such as the EORTC-QLQ C30, FACT/GOG-Ntx, HADS, and MFI-20, which capture fatigue, neuropathy, anxiety, depression, and overall well-being [36, 37].

While numerous validated tools exist, they typically address nutrition, physical activity, or quality of life separately [38]. Given evidence that interventions in these domains can act synergistically [27], a combined assessment appears beneficial. The INTEGRATION study (DRKS00020208) therefore introduced the Resource-Oriented Needs Assessment (RoBa) and its corresponding RoBa-Score, designed to capture multimodal supportive care needs in oncology.

This study aims therefore to evaluate the feasibility, applicability, and validity of this combined instrument compared with its individual components, including subgroup analyses in gastrointestinal (GI) and non-GI tumor patients.

## Materials and Methods

### Composition of the multimodal resource-oriented needs analysis (RoBa) and its derived scoring system

The multidimensional RoBa assessment was assembled from twelve published and validated single assessments spanning three domains—physical activity, nutrition and quality of life— with four assessments per domain to ensure balanced weighting (**Table 1**). Variables were selected based on contemporary evidence for diagnostic and prognostic value and on their modifiability through exercise and/or nutrition therapy. The RoBa integrates standardized questionnaires alongside objectively measurable parameters (device-based measures, laboratory markers, and clinical data). <u>Physical activity domain:</u> strength (handgrip dynamometry [38]); endurance (cardiopulmonary exercise testing [CPET] [39] using a 30/15 protocol; 30/15 Intermittent Fitness Test); coordination (one-leg stance test [40]); and habitual physical activity behavior over seven days (accelerometry [41]). <u>Nutrition status domain</u>: body-mass-index (BMI [42]); body composition dervived from bioelectrical impedance analysis (BIA [43, 44]); inflammatory/metabolic status determined by the modified Glasgow-Prognostic-Score (mGPS [45]), and signs of malnutrition (PG-SGA [46]). <u>Quality-of-life domain.</u> fatigue (MFI-20 [47]); chemotherapy-induced peripheral neuropathy (FACT/GOG-Ntx [48]); anxiety and depression (HADS [49]); and health-related quality of life (EORTC-QLQ-C30 [50]) (**Table 1**).

**Table 1:** Composition of the Multimodal Resource-oriented Needs Assessment Score (RoBa Score)

| RoBa Domain | Parameter | Assessment | Domain Score | Reference |
| --- | --- | --- | --- | --- |
| Physical Activity | Strength | Handgrip [kg] | 0: >25th percentile<br>1: >10th - ≤25th percentile<br>2: ≤10th percentile<br>per age and gender based on a norm cohort | (1) |
|  | Endurance | Spiroergometry [VO<sub>2max</sub>] | 0: fair<br>1: poor<br>2: very poor<br>per age and gender based on a norm cohort | (2) |
|  | Balance | Unipedal stance [sec] | 0: >Mean-1xSE<br>1: >Mean-2xSE and ≤Mean-1xSE<br>2: ≤Mean-2xSE<br>per age and gender based on a norm cohort | (3) |
|  | Physical activity behavior | Accelerometry [Time spent in MVPA min/week] | 0: >300 min/week<br>1: 150-300 min/week<br>2: <150 min/week | (4) |
| Nutrition | Anthropometry | Body Mass Index [kg/m<sup>2</sup>] | 0: ≥20.0 – 24.9<br>1: ≥25.0 OR <20.0 <u>without</u> weight loss <sup>(1)</sup><br>2: <18.5 OR <20.0 kg/m<sup>2</sup> <u>with</u> weight loss <sup>(1)</sup> | (5) |
|  | Body composition | Bioelectrical impedance analysis [Phase angle] | 0: >Mean-1xSD<br>1: ≤Mean-1xSD and >Mean-2xSD<br>2: <Mean-2xSD<br>per age and gender based on a norm cohort | (6, 7) |
|  | Inflammation and metabolism | mGPS [Serum CRP mg/l & Albumin g/l] | 0: CRP ≤10 and Albumin ≥35<br>1: CRP >10 and Albumin ≥35<br>2: CRP >10 and Albumin <35 | (8) |
|  | Nutritional status | PG-SGA (questionnaire) | 0: A (well nourished)<br>1: B (moderately malnourished/suspected malnutrition)<br>2: C (severely malnourished) | (9) |
| Quality of Life | Fatigue | MFI-20 (questionnaire) | 0: <75<sup>th</sup> percentile<br>1: ≥75<sup>th</sup> percentile and <90<sup>th</sup> percentile<br>2: ≥90<sup>th</sup> percentile<br>per age and gender based on a norm cohort | (10) |
|  | Therapy induced polyneuropathy | FACT/GOG-Ntx (questionnaire) | 0: 0-11 (item score)<br>1: 12-22 (item score)<br>2: >22 (item score) | (11) |
| | Anxiety and depression | HADS (questionnaire) | 0: 0-13<br>1: 14-21<br>2: $\geq 21$ | (12) |
| | Quality of life | EORTC QLQ-C30 (questionnaire) | 0: $>\text{Mean}-1\text{xSD}$<br>1: $\leq\text{Mean}-1\text{xSD}$ and $>\text{Mean}-2\text{xSD}$<br>2: $<\text{Mean}-2\text{xSD}$<br>per age and gender based on a norm cohort | (13) |

**Legend to Table 1:** Twelve individual validated assessments are included in the RoBa score. Four assessments are grouped and form one of three RoBa domains, i.e. physical activity, nutrition, quality of life status. Abbreviations: BMI=Body Mass Index; PG-SGA=Patient-Generated Subjective Global Assessment; MFI=Multidimensional Fatigue Inventor; FACT/GOG-Ntx=Functional Assessment of Cancer Therapy/Gynecologic Oncology Group—Neurotoxicity; HADS= Hospital anxiety and depression scale; EORTC=European Organization for Research and Treatment of Cancer Core Quality of Life Questionnaire; MVPA=Moderate to Vigorous Physical Activity.
<sup>(1)</sup> Significant underweight is defined as a BMI $<18.5\text{kg/m}^2$ , or BMI $<20\text{kg/m}^2$ AND unintentional weight loss of $>5\%$ of baseline weight in the previous 3-6 months.

Each single assessment was mapped to a 3-level scale—0 = no need/deficit, 1 = moderate need/deficit, 2 = high need/deficit—according to the instrument’s original publication, internationally accepted reference values, and relevant guidelines. Within each domain, single-assessment scores were summed to yield a domain score (“domain score”); domain scores were then summed to a total RoBa score. With twelve single assessments each scored 0–2, the maximum total score is 24.

#### Physical activity domain

Strength (handgrip). Isometric maximal grip strength was measured with a digital Jamar dynamometer; results reported in kilograms. Categorization used age- and sex-specific reference values [39, 40]: 0 = > 25th percentile; 1 = > 10th to ≤ 25th percentile; 2 = ≤ 10th percentile. Endurance (CPET, 30/15 protocol). Aerobic capacity and relative VO_2_max were assessed by spiroergometry. Breath-by-breath analysis quantified VO_2_, VCO_2_, minute ventilation (VE), and breathing frequency. With progressive workloads, VO_2_ increases linearly until steady-state; beyond steady-state, VO_2_max reflects maximal oxygen transport (mL·min_−1_). Categorization followed age- and sex-normalized strata [41]: 0 = fair or better; 1 = poor; 2 = very poor. Coordination (one-leg stance). The longest stable single-leg stance from three trials with eyes open and closed (maximum 45 s) was recorded. Categorization was based on a normative reference sample stratified by age and sex [42]: 0 = > mean – 1 SD; 1 = ≤ mean – 1 SD to > mean – 2 SD; 2 = ≤ mean – 2 SD. Because of the 45-s ceiling, negative thresholds arose in some age groups; therefore, the 50–59-year group served as the reference for all participants. Habitual physical activity (accelerometry). Participants wore a triaxial accelerometer (Move4; 100 Hz, Movisens GmbH) continously for 7 consecutive days. The device recorded frequency, duration, and intensity of physical activity and estimates activity-induced energy expenditure. Data were analyzed using the Movisens DataAnalyzer (version 1.15.1, module base and energy expenditure), which calculates time spent in moderate to vigorous physical activity (mvpa) based on pre-defined MET-/h ranges. Categorization was based on total time spent in mvpa over one week and WHO recommendation thresholds [32]: 0 = > 300 min/week; 1 = 150-300 min/week; 2 = < 150 min/week.

#### Nutrition domain

BMI was calculated on current weight and height, and the RoBa score was categorized 0 = ≥ 20.0–24.9 kg/m^2^; 1 = ≥ 25.0 kg/m^2^ or 18.5–< 20.0 kg/m^2^ without unintentional 3–6-month weight loss of > 5%; 2 = < 18.5 kg/m^2^ or < 20.0 kg/m^2^ with > 5% unintentional weight loss in the prior 3–6 months. For BIA, a phase-sensitive, multi-frequency BIA device was used (BIACORPUS RX 4000 (MediCAL), software BodyComp V9.0). BIA measurements were performed at 50 kHz in resting patients in a supine position during the morning hours and after overnight-fasting and with an emptied urine bladder. The phase angle (PA) was taken as a direct measure of captures time shift between current and voltage maxima and indexes body cell mass. Categorization was based on a normative reference sample stratified by age and sex. 0 = > mean – 1 SD; 1 = ≤ mean – 1 SD to > mean – 2 SD; 2 = ≤ mean – 2 SD. Modified Glasgow Prognostic Score (mGPS) [29]. CRP and albumin were extracted from routine clinical blood draws within ± 1 week of the RoBa time point. Categorization [56]: 0 = CRP ≤ 10 mg/L and albumin ≥ 35 g/L or CRP ≤ 10 mg/L with albumin < 35 g/L; 1 = CRP > 10 mg/L and albumin ≥ 35 g/L; 2 = CRP > 10 mg/L and albumin < 35 g/L. Nutritional status (PG-SGA) [43]. The PG-SGA combines patient-reported and professional components to evaluate weight, intake, symptoms, functional and disease status, metabolic stress, and physical examination (subcutaneous fat, muscle mass, edema), and considers age, comorbidities, disease stage, fever, and corticosteroid use [28]. The Global Assessment classifies: A (well nourished), B (moderate/suspected malnutrition), C (severe malnutrition). RoBa categorization: 0 = A; 1 = B; 2 = C.

#### Quality-of-life domain

Fatigue (MFI-20). The 20-item MFI-20 assesses fatigue over the past week across five subscales (general, physical, mental fatigue; reduced activity; reduced motivation) [49]. Items use a 5-point Likert scale; subscales (max 20 each) sum to a total (max 100; lower scores = less fatigue). Categorization was based on a normative reference sample fort he general fatigue subscale stratified by age and sex [49]: 0 = < 75th percentile; 1 = ≥ 75th percentile and < 90th percentile; 2 = > 90th percentile. Chemotherapy-induced peripheral neuropathy (FACT/GOG-Ntx). The FACT/G includes 27 core items; the Ntx subscale adds 11 items (0 = “not at all” to 4 = “very much”), summed to a maximum of 44 (lower scores = less CIPN). Categorization [50, 51]: 0 = 0–11; 1 = 12–22; 2 = > 22. Anxiety and depression (HADS). HADS comprises 14 items, forming anxiety (A) and depression (D) subscales (0–21 each; total 0–42; higher scores indicate greater symptom burden). Subscale scores ≥ 11 indicate caseness. RoBa categorization [52]: 0 = 0–13; 1 = 14–20; 2 = > 20. Health-related quality of life (EORTC-QLQ-C30). The core questionnaire captures the previous week across five functional and nine symptom subscales [57]; optional diagnosis/treatment modules can be added [58]. Items are transformed to 0–100; higher functional scores denote better functioning, whereas higher symptom scores denote worse symptoms; therefore, subscales were analyzed individually (not as a single total). Only the Global Health/Quality of Life subscale was included for The study, based on a normative reference sample stratified by age and sex [53, 59]: 0 = > mean – 1 SD; 1 = ≤ mean – 1 SD to > mean – 2 SD; 2 = ≤ mean – 2 SD.

### Study design and ethical statement

The study was conducted as a prospective, non-interventional, single arm trial at two German Comprehensive Cancer Centers: the Comprehensive Cancer Center Munich, LMU Munich (CCCM) and the Center for Integrated Oncology (CIO), University Hospital Cologne. The study protocol followed ethical considerations stated by the Declaration of Helsinki, and ethical approval was obtained from the institutional review board (LMU Munich, Project No. 21-1027). From all included patients written informed consent was obtained.

### Study population

All consecutive cancer patients at CCCM and CIO were screened.

<u>Inclusion criteria</u>

- Diagnosis of cancer
- ECOG performance status ≤ 2
- Age ≥ 18 years
- Written informed consent

<u>Exclusion criteria</u>

- Contraindications to perform physical activity measurements (spiroergometry), i.e. including: severe heart failure (NYHA III–IV); therapy-refractory arterial hypertension grade III–IV; global respiratory failure, COPD > III°; therapy-refractory thrombocytopenia < 10,000/μL; clinically relevant uncontrolled hemorrhagic diathesis; unstable coronary syndrome; unstable arrhythmias; uncontrolled cerebral seizure disorder
- Contraindications to BIA: implanted pacemaker or defibrillator; non-removable active prostheses with electronic control
- Pregnancy

### Endpoints

Primary endpoint: Feasibility and validity of the RoBa score with its twelve single assessments in a clinical setting.

Secondary endpoints:

- Agreement between needs classification based on validated single-assessment scoring and the corresponding classification using RoBa-specific cut-points.
- Comparison of needs classification derived from the set of single assessments with the classification based on the RoBa total score.
- Subgroup analyses by tumor entity (gastrointestinal [GI] vs. non-GI).

### Study procedures and RoBa measurements

After informed consent, a study check-in recorded brief medical history (recent weight change; acute symptoms such as nausea, neuropathy, pain; infection signs) and vital signs (blood pressure, pulse, respiratory rate). The check-in also screened for contraindications to undergo specific measurements, i.e. spiroergometry, BIA.

On Day one, all RoBa assessments were completed except accelerometry. At the end of Day 1, participants received an accelerometer, wore it continuously for seven consecutive days, and returned it on Day 8.

### Statistical analysis

Demographics were summarized and analyzed descriptively: categorical variables as counts and percentages; continuous variables as mean, standard deviation, and quartiles. Group comparisons used t-tests, Mann–Whitney U tests, or χ^2^ tests (Fisher’s exact test, as appropriate). Interactions of RoBa domain and total score results were performed with Spearmann correlation analyses. Statistical significance was defined as p<0.05.

## Results

### Clinical characteristics

Of 369 screened patients, 62 cancer patients (36 female, 26 male) were enrolled, and baseline clinical characteristics and cancer entities are shown in **Table 2 and 3**. The most common comorbidities were nutritional and metabolic disorders such as thyroid disease, diabetes mellitus, or dyslipidemia (38.7%). Most participants (69.4%) had been diagnosed within the previous year. The most common therapy-related complaints were non-specific symptoms (e.g., dry skin, muscle weakness, dizziness; 62.9%), neurological changes (taste alterations, chemotherapy-induced neuropathy; 58.1%), general symptoms (fatigue, nausea, anorexia; 45.2%), and gastrointestinal complaints (diarrhea, constipation; 45.2%). Of 62 participants, 25 had GI tumors and 37 non-GI tumors. Female sex was less frequent in the GI group (40% vs. 70.3%, p = 0.018). Nutrition-related side effects were more common in GI patients (48% vs. 21.6%, p = 0.029).

**Table 2:** Clinical baseline characteristics of study participants.

| Characteristics | All (n=62) | GI (n=25) | Non-GI (n=37) | p-value |
| --- | --- | --- | --- | --- |
| Age (years), Median (Range) | 56 (27–81) | 59 (42–78) | 55 (27–81) | 0.073 |
| Sex, n (%) |  |  |  |  |
| Female | 36 (58.1) | 10 (40.0) | 26 (70.3) | **0.018** |
| Male | 26 (41.9) | 15 (60.0) | 11 (29.7) |  |
| ECOG, n (%) |  |  |  |  |
| 0 | 47 (75.8) | 20 (80.0) | 27 (73.0) | 0.526 |
| 1 | 15 (24.2) | 5 (20.0) | 10 (27.0) |  |
| Anthropometrics, Median (Range) |  |  |  |  |
| Height (cm) | 169 (143–193) | 174 (158–193) | 165 (143–185) | **0.009** |
| Weight (kg) | 69.6 (44.6–135.4) | 69.7 (51.7–91.3) | 69 (44.6–135.4) | 0.840 |
| Waist circumference (cm) | 88 (58–137) | 87 (67–103) | 88 (58–137) | 0.640 |
| Comorbidities, n (%) |  |  |  |  |
| Nutritional or metabolic diseases* | 24 (38.7) | 8 (32.0) | 16 (43.2) | 0.373 |
| Cardiovascular diseases | 10 (16.1) | 5 (20.0) | 5 (13.5) | 0.496 |
| Musculoskeletal diseases | 8 (12.9) | 4 (16.0) | 4 (10.8) | 0.550 |
| Allergies and intolerances | 13 (21.0) | 6 (24.0) | 7 (18.9) | 0.630 |
| Neurological and psychiatric diseases | 9 (14.5) | 2 (8.0) | 7 (18.9) | 0.231 |
| Other secondary diagnoses** | 23 (37.0) | 10 (40.0) | 13 (35.1) | 0.785 |
| No secondary diagnoses | 16 (25.8) | 8 (32.0) | 8 (21.6) | 0.360 |

**Legend to Table 2:** Clinical baseline characteristics of study participants, entire cohort and GI vs. non-GI cancer subgroups; GI: gastrointestinal tumors, non-GI: all other entities; bold print: statistically significant ( $p < 0.05$ ) \* Thyroid disease, diabetes mellitus, elevated blood lipids or cholesterol or triglycerides, gout or uric acid disorder, congenital metabolic disorder. \*\* Skin disease(s), kidney disease(s), digestive system disease(s), lung disease(s), infectious diseases.

**Table 3:** Cancer Diagnosis Distribution.

| **Cancer Types** | **Number (total, relative)** |
| --- | --- |
| **GI (n=25, 40.3%)** |  |
| Colorectal cancer | 10 (16.1) |
| Pancreatic cancer | 9 (14.5) |
| Other* | 6 (9.6) |
| **Non-GI (n=37, 59.7%)** |  |
| Breast cancer | 15 (24.2) |
| Hematologic neoplasia | 10 (16.1) |
| Other** | 12 (19.4) |

**Legend to Table 3:** \* Includes: Esophagus (C15), Small intestine (C17), Liver and intrahepatic bile ducts (C22), Other and unspecified parts of the biliary tract (C24). \*\* Includes: Female genital organs (C51–C58), Respiratory and other intrathoracic organs (C30–C39), Melanoma and other malignant neoplasms of skin (C43–C44), Male genital organs (C60–C63), Urinary organs (C64–C68), Malignant neoplasms of ill-defined, secondary, and unspecified sites (C76–C80).

### Single-assessment results

#### Physical activity status

Median handgrip strength was 34.4 kg; no participant scored 2. Median one-leg stance was 45 s (maximum), yielding a score of 0 in 87.1%. Median relative VO_2_max was 25.7 ml/kg/min; scores were distributed as 45.2% = 0, 24.2% = 1, 27.4% = 2. Median mvpa was 352; most participants scored 1 (n=37) (**Figure 1**). Given age and sex differences in handgrip, the median 34.4 kg is context-dependent; relative to reference means at ∼56 years (men 46.2 kg; women 27.5 kg) [40], the cohort median sits between, slightly closer to the female mean— consistent with the sex distribution. No participant reached handgrip score 2; most scored 0 (54/62), suggesting either the cohort did not span the full RoBa range. One-leg stance performance was high (median 45 s; ceiling), exceeding general-population norms (mean 33.4 s) [42], and yielding predominantly score 0 (87.1%). Because published age/sex norms can produce negative cut-offs when capped at 45 s, this study used the 50–59-year reference as a pragmatic solution; age/sex-specific norms from the original source remain preferable where applicable. Median relative VO_2_max was 25.7 ml/kg/min. Using ∼56 years as a reference, this maps to “fair” in women (score 0) and “very poor” in men (score 2) per Heyward et al. [41]. Overall, 51.6% had elevated need (score 1 or 2), consistent with evidence that targeted endurance training can improve CRF during active treatment across entities [92]. Median mvpa (352) exceeded WHO recommendations (150–300 min/week) [93], which is notable given expected activity reductions during cancer therapy; high education may have contributed [95]. However, mapping accelerometer-derived mvpa to questionnaire-derived MET-h/wk is methodologically problematic, as no validated conversion exists and definitions of mvpa vary [32–34, 96–100]. A more defensible RoBa approach would be to anchor accelerometer data to WHO thresholds [101], which yields a distribution more consistent with other activity assessments.

**Figure 1:**
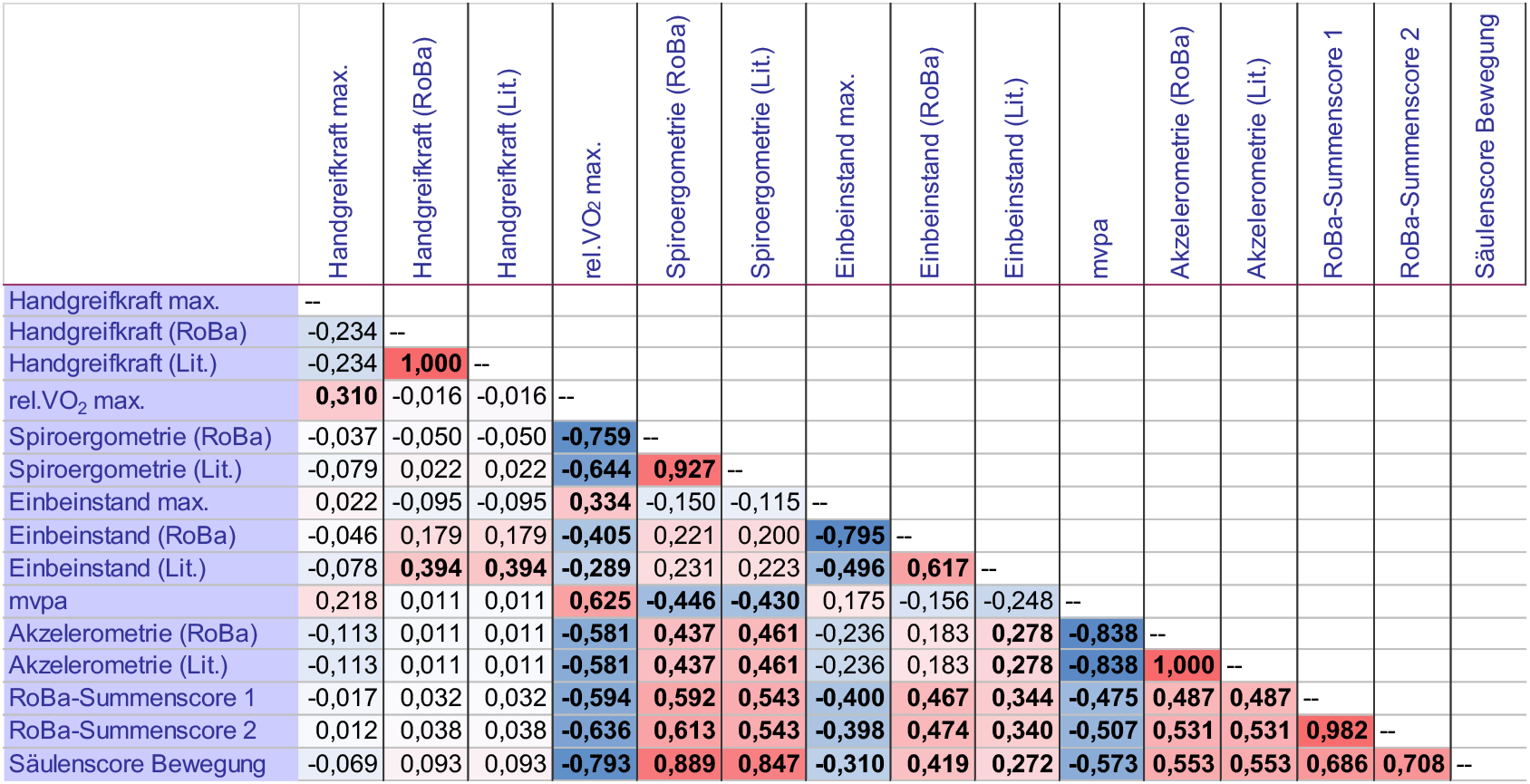
Physical Fitness Status – Correlation Analyses. Spearman correlation coefficients for the exercise scores; bold = statistically significant (p < 0.05); Color scale: dark blue: > −0.6, strong negative correlation; light blue: < −0.3, weak negative correlation; deep red: > 0.6, strong positive correlation; light red: < 0.3, weak positive correlation. WHtR = waist-to-height ratio; BIA = bioelectrical impedance analysis; CRP = C-reactive protein; mGPS = modified Glasgow Prognostic Score; PG-SGA = Scored Patient-Generated Subjective Global Assessment; RoBa = resource-oriented needs assessment score; Lit. = evaluation according to original literature source; Column score Nutrition 1: with BMI; Column score Nutrition 2: with WHtR; RoBa total score 1: formed with column score Nutrition 1; RoBa total score 2: formed with column score Nutrition 2.

#### Nutrition status

Median BMI was 24.1 kg/m^2^. Median phase angle was 5.4° (range 2.8–7.2). Distributions of the categorized single nutrition assessments are shown in **Figure 2**. Albumin median was 42.0 g/L; CRP median 10.0 mg/L. Most patients (n=45) had an mGPS of 0. By PG-SGA, 51.6% were classified A (well-nourished), 46.8% B (moderate malnutrition), and one patient C (severe malnutrition). The cohort’s median BMI (24.1 kg/m^2^) fell in the WHO normal range [44] and was comparable to other ambulatory oncology cohorts (e.g., mean BMI 22.8 kg/m^2^) [80].

**Figure 2:**
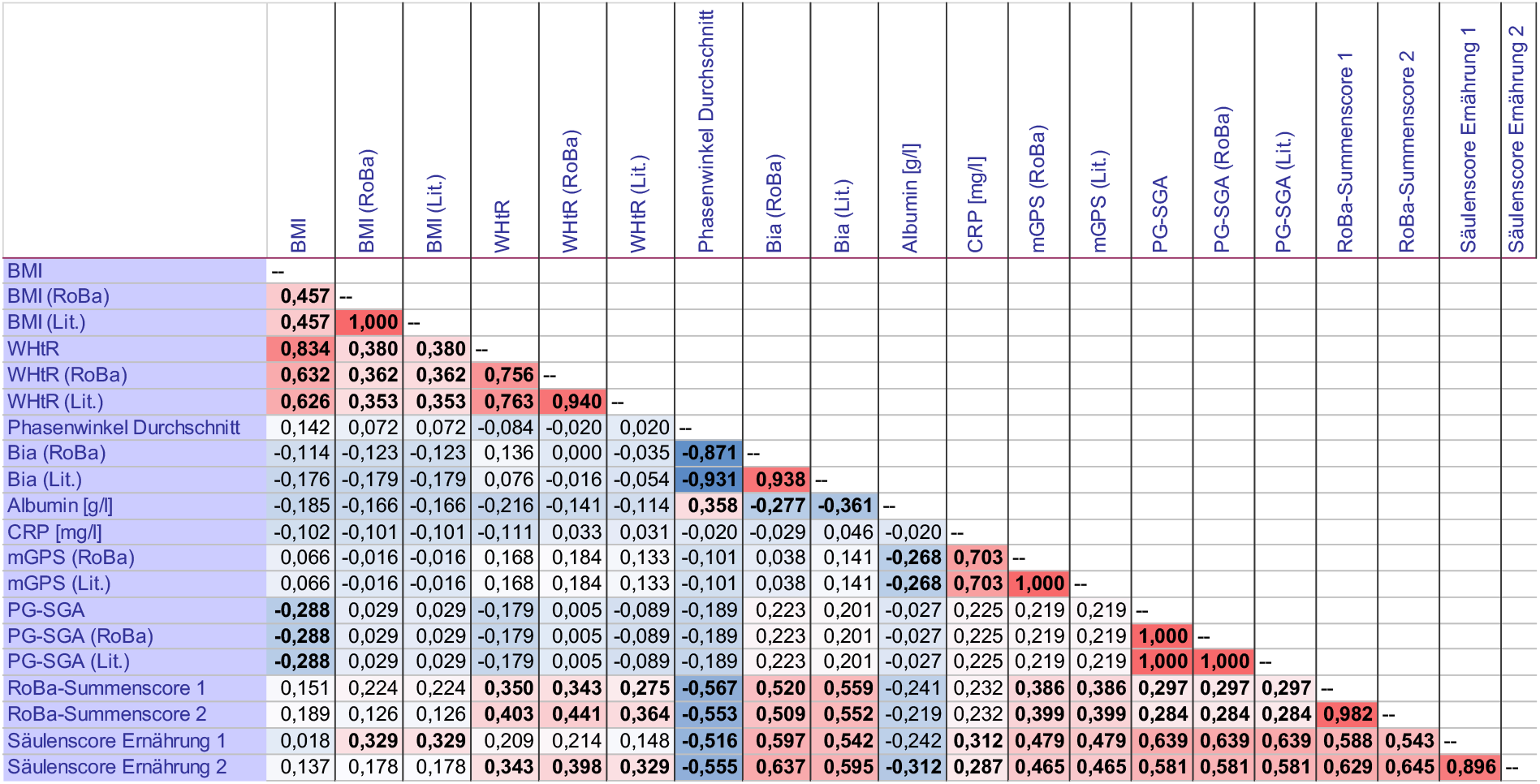
Nutrition Status – Correlation Analyses. Spearman correlations for the nutrition scores; bold = statistically significant (p < 0.05). Color scale: dark blue: > −0.6, strong negative correlation; light blue: < − 0.3, weak negative correlation; deep red: > 0.6, strong positive correlation; light red: < 0.3, weak positive correlation. WHtR = waist-to-height ratio; BIA = bioelectrical impedance analysis; CRP = C-reactive protein; mGPS = modified Glasgow Prognostic Score; PG-SGA = Scored Patient-Generated Subjective Global Assessment; RoBa = resource-oriented needs assessment score; Lit. = evaluation according to original literature source; Column score Nutrition 1: with BMI; Column score Nutrition 2: with WHtR; RoBa total score 1: formed with column score Nutrition 1; RoBa total score Nutrition 2: formed with column score Nutrition 2.

Relative to Munich and Bavaria population data (24.7 vs. 25.8 kg/m^2^), the cohort appears relatively lean [81]. Phase angle (median 5.4°, range 2.8–7.2°) was similar to baseline values reported in breast cancer cohorts [83]. Notably, BIA (RoBa) classified more participants as impaired (19 with score 2) than mGPS, PG-SGA, or anthropometry, suggesting that BIA—by capturing cellular integrity and body cell mass—may detect early nutritional compromise not yet reflected in anthropometric indices. Median albumin (42.0 g/L) was normal, whereas median CRP (10.0 mg/L) was elevated relative to institutional reference ranges, consistent with systemic inflammation in cancer [84]. Most participants had mGPS 0, indicating adequate protein status and no systemic inflammation. PG-SGA categories (A 51.6%, B 46.8%, C 1.6%) were comparable to prior oncology cohorts [80, 85].

#### Quality of life status

Median scores were: MFI-20 General Fatigue = 11, FACT/GOG-Ntx = 7.5, HADS = 8.0, and EORTC QLQ-C30 Global Health = 66.7. As shown in **Figure 3**, most participants scored 0 across all QoL single assessments. Median MFI-20 General Fatigue (11) suggests greater fatigue than healthy norms (men 8.0; women 8.7) [49]—expected in oncology—yet less than in some cancer cohorts (means 15.8–19.9) [103], consistent with most patients scoring 0.

**Figure 3:**
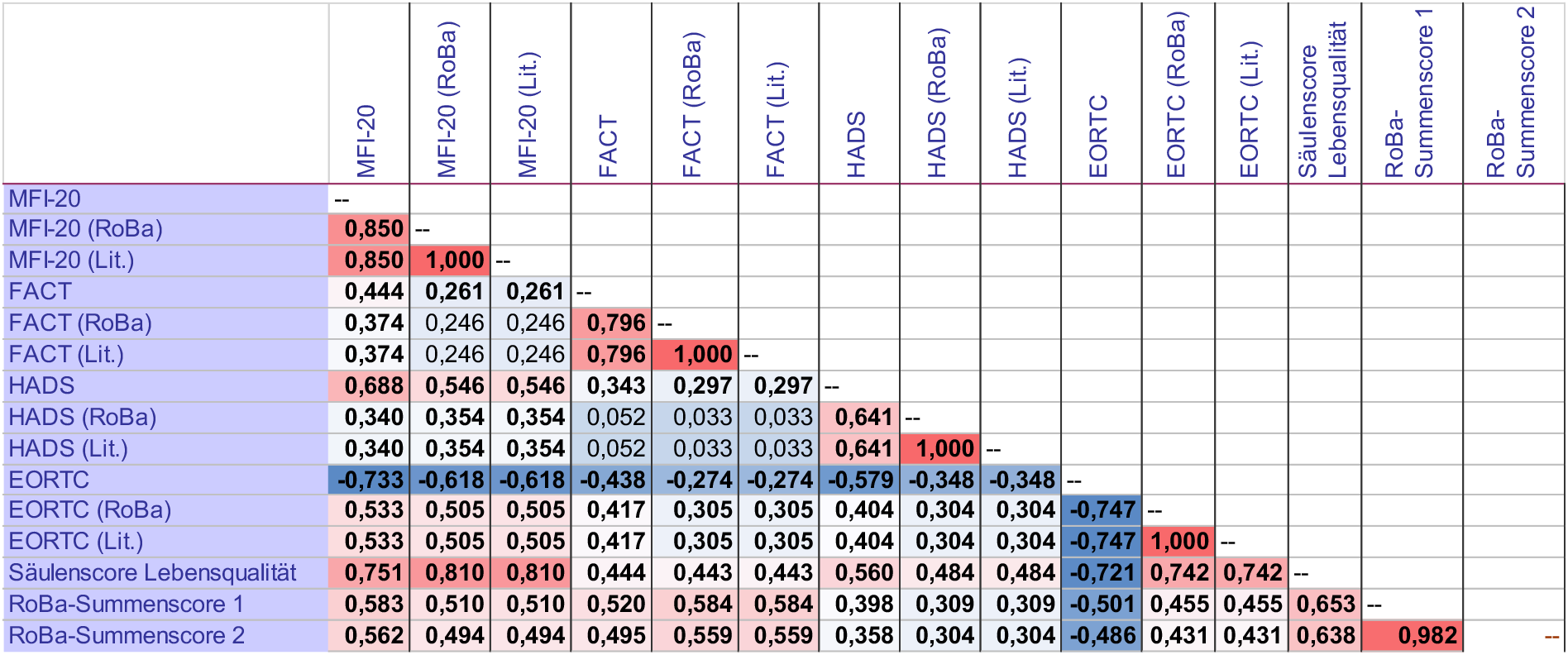
Quality of Life – Correlation Analyses. Spearman correlation coefficients for the quality-of-life scores; bold = statistically significant (p < 0.05); Color scale: dark blue: > −0.6, strong negative correlation; light blue: < −0.3, weak negative correlation; deep red: > 0.6, strong positive correlation; light red: < 0.3, weak positive correlation. MFI-20 = Multidimensional Fatigue Inventory; FACT = Functional Assessment of Cancer Therapy / Gynecologic Oncology Group – Neurotoxicity; HADS = Hospital Anxiety and Depression Scale; EORTC = European Organization for Research and Treatment of Cancer; RoBa = resource-oriented needs assessment score; Lit. = evaluation according to original literature source; RoBa total score 1: formed with column score Nutrition 1 or BMI; RoBa total score 2: formed with column score Nutrition 2 or WHtR (waist-to-height ratio).

FACT/GOG-Ntx (median 7.5) was well below values seen in cohorts with moderate–severe CIPN [50]. HADS (median 8.0) was lower than in metastatic cohorts on active treatment [104]. EORTC QLQ-C30 Global Health (median 66.7) approximated the European population mean (66.1) [53] and was higher than values reported pre-exercise intervention in breast cancer survivors [106].

### Correlation analyses and alignment of RoBa scoring

All domain scores correlated strongly and comparably with total scores (r = 0.543–0.708), supporting balanced weighting. In nutrition, BIA often yielded higher need than other measures, consistent with its sensitivity to early cellular changes under oncologic therapy [117, 118]; phase angle’s inverse correlations with domain and total scores align with its prognostic value across settings [118, 119].

In the activity domain, CPET/VO_2_max showed the strongest agreement with the domain score (r = 0.889) and meaningful links to total scores, consistent with CPET’s status as the CRF gold standard [35, 120]. Accelerometry showed the most negative deviations from the domain score, reinforcing the need for domain-appropriate thresholds (e.g., WHO) in RoBa. Handgrip demonstrated limited spread (no score 2) and few correlations, suggesting cut-points may need refinement; its primary clinical utility may relate more to frailty detection than to CRF per se [39, 118].

All QoL instruments correlated robustly with the QoL domain and total scores; strong interrelations among fatigue, HADS, and EORTC replicate prior oncology findings [123, 124].

### Subgroup analyses − GI vs. non-GI cancer patients

Women were more frequent in the non-GI group, reflecting the high proportion of breast cancer. GI patients more often reported weight/nutrition-related side effects, consistent with prior data [125]. Otherwise, baseline characteristics were balanced, supporting comparability and arguing against selection bias.

Among all single assessments across domains, only PG-SGA differed significantly between groups, indicating worse nutritional status in GI patients—consistent with the high malnutrition risk in GI cancers [126–128]. The absence of differences in other nutrition metrics may reflect the short interval since diagnosis for many participants and/or limited sample size.

Only Nutrition Domain Score 2 (WHtR-based) differed by group, with more non-GI patients scoring 0 (13 vs. 2), reinforcing PG-SGA findings and literature indicating lower nutrition-support needs in non-GI entities [126–128]. No group differences emerged in total scores or in deviations between single and domain scores, except for WHtR vs. Nutrition Domain Score 2. This may indicate entity-independent classification performance—or limited power.

Fewer significant correlations were observed in GI than in non-GI, particularly in the nutrition domain. Nonetheless, the strongest relationships were preserved across subgroups: inverse associations of phase angle with nutrition/total scores, strong CPET/VO_2_max correlations with the activity domain, and consistent QoL intercorrelations and links to total scores.

## Discussion

In this study, we developed and evaluated a multidimensional assessment tool based on twelve validated single assessments to comprehensively characterize the supportive care needs and resources of cancer patients. We observed that testing cancer patients with this comprehensive assessment battery was feasible. Moreover, the scoring system established with the RoBa score showed overall strong correlations of the individual assessments with the respective domain score and the total RoBa score.

The study cohort comprised 62 patients, and women were slightly overrepresented (58.1% vs. 41.9%), a pattern consistent with previous and published studies in supportive care medicine. Patients with poorer performance status (ECOG = 2) were markedly underrepresented although we made efforts to especially include also patients of lower clinical performance status. Although permitted per inclusion criteria and actively screened, a lot of those patients were not willing to participate for medical or personal reasons (e.g., concurrent radiochemotherapy, high visit burden, perceived lack of fitness, travel distance, or the length of the RoBa visit). Similar underrepresentation of ECOG 2 (or Karnofsky 50–60%) is reported in other nutrition/physical-activity studies [21, 66, 67], though some cohorts have enrolled ECOG 2 patients [68–70].

To capture the full range of nutritional, activity, and QoL needs, inclusion criteria emphasized gastrointestinal entities—particularly pancreatic and colorectal cancers—which was achieved (40.3% of participants). Breast cancer accounted for 24.2%, mirroring many lifestyle studies [65]. High participation among women with breast cancer likely reflects comparatively favorable survival [71] and robust evidence linking healthy lifestyle to improved outcomes and survival [72–74]. Many of the included patients reported ongoing treatment-related adverse effects—most commonly neurological changes, non-specific/general symptoms, and gastrointestinal complaints—consistent with known toxicity profiles that vary by agent, dose, and route of administration. All included participants completed the RoBa assessments as planned. Missing data were not due to non-feasibility but to technical issues.

Differences between RoBa categorization and original references were most pronounced for phase angle (BIA) and CPET because original sources use six categories, whereas RoBa collapses to three (**Table 1**). For phase angle, 59.7% of classifications differed. While granularity is greater with six categories, revisiting cut-points may improve alignment, particularly as BIA was the only nutrition assessment not dominated by score 0. For CPET, differences in 15 cases largely reflected consolidation of higher-fitness categories (“superior/excellent/good”) into score 0; retaining RoBa’s 3-tier scheme seems reasonable, optionally annotating the additional high-fitness strata as sublevels of 0. For one-leg stance, eight discrepancies arose; age/sex-specific norms from the source [42] should be preferred in future RoBa implementations where feasible.

Limitations of this study comprise of the small sample size and the lack of patients with a poor performance status, i.e. ECOG 2 and higher, that particularly represent a group of patients with high needs of supportive care. However, in this pilot study we found the RoBa multidimensional assessment being feasible in clinical practice, and informative in delineating domain-specific needs, particularly highlighting nutritional support requirements. Methodological refinements—most notably (i) adopting WHO-anchored thresholds for accelerometry, (ii) revisiting handgrip cut-points, and (iii) considering more granular BIA and CPET categorization—may enhance sensitivity and alignment with clinical reality. Future studies should aim to particularly include patients with lower clinical performance status (i.e. ECOG = 2-3) and expand homogenous subgroup sizes to validate the RoBa score’s generalizability and to optimize thresholds for clinical decision-making.

## Data Availability

Major data produced in the present work are contained in the manuscript. Additional data are available upon reasonable request to the authors.

## Acknowledgements

This study was in part funded by the Innovationsfond of the Joint Federal Committee (Gemeinsamer Bundesausschuss, GBA), Germany (Project number 01NVF18018). We are also grateful for the institutional support of the German Society of Nutritional Medicine (DGEM), the German Association for Excercise Science and Therapy (DVGS), the department of Molecular and Cellular Excercise Sciences at the German Sport University Cologne, and the Nutrition, Metabolism and Excercise Working Group of the German Society of Hematology and Oncology (DHGO).

## Author contributions

EK, ST and FTB developed the study and wrote the protocol. EK and AT included patients and obtained primary study data. EK, ST and CW analyzed the data. ST and EK wrote the manuscript draft. TN, NE, HJB, NZ and ML contributed to the trial concept and the manuscript draft. All authors reviewed the manuscript.

